# The effect of family history and polygenic risk scores on general and abdominal obesity in the Lifelines Cohort Study

**DOI:** 10.1101/2025.09.22.25336364

**Authors:** Rujia Wang, Catharina A. Hartman, Lifelines Cohort Study, Peter M. Visscher, Harold Snieder

## Abstract

**Background:** Polygenic risk scores (PRSs) have recently become popular in predicting genetic susceptibility to common complex diseases. However, as PRSs only capture part of the genetic risk, imperfect prediction of outcomes is expected. Family history may capture additional genetic effects as well as those of shared family environment. Within-family design have the advantage to control potential bias, such as gene-environmental correlation, assortative mating, and population stratification. Our aims are to investigate the relative influence of PRS and family history on general and abdominal obesity using between and within family approaches.

**Methods:** We included 50,747 participants of the Lifelines Cohort Study. Body mass index (BMI) and waist-hip-ratio (WHR) were calculated by measured height, weight, waist circumference (WC) and hip circumference. Family history was defined as the number of obese first-degree relatives. PRSs were calculated based on recent large genome-wide association studies for BMI, WC and WHR. The between-family PRS was defined as the mean sibling PRS, and the within-family PRS as the difference between the individual and mean PRS. Linear mixed regression models were used to estimate the effect of (between- and within-family) PRS and family history on BMI, WC and WHR. Structural equation modeling was used to estimate the relative effects of parental PRS (via offspring’s PRS) and parental phenotype on offspring’s phenotype.

**Results:** PRS and family history together explained 11.81%, 7.26% and 3.62% of the variance in BMI, WC and WHR, respectively, where family history explained additional variance on top of PRS. Among 2,003 parent-offspring trios, transmission of the PRSs from parents to offspring (*β*=0.48-0.52) resulted in a significant effect of offspring’s PRSs on offspring’s BMI, WC and WHR (*β*=0.14-0.16). In addition, parental BMI, WC and WHR, were strongly positively associated with offspring’s BMI, WC and WHR (*β*=0.14-0.27), independent of the parent-offspring transmission. Between- and within-family PRS had similar effect sizes, while between-family PRS explained more variance than within-family PRS for obesity indices among siblings (1.91% vs 6.70% for BMI, 1.25% vs 4.10% for WC, and 0.72% vs 2.61% for WHR).

**Conclusions:** The combination of PRS and family history improved the prediction of indices for general and abdominal obesity. In addition to the genetic effects captured by the parent-offspring PRS transmission, parental BMI, WC and WHR also independently influenced offspring’s BMI, WC and WHR, reflecting additional genetic effects not captured by PRS as well as effects of the shared family environment. Similar effects of between- and within-family PRSs indicated little bias (e.g., gene-environmental correlations, assortative mating, and population stratification) in the genetic effects on indices of obesity.

## INTRODUCTION

The prevalence of obesity has increased worldwide in recent decades, with estimates of 15% among the world’s adult population in 2020^1^. General obesity is defined by a body mass index (BMI) of 30 kg/m^2^ and higher and is a multifactorial disease, with complex pathogenesis related to genetics^2^, socioeconomic status^3^, and lifestyles (e.g., dietary habits and physical activity)^4,5^. High BMI is a risk factor for the incidence and development of cardiovascular disease^6,7^. In 2017, high BMI caused 4.7 million deaths globally^8^. Abdominal obesity, that can be defined by waist circumference (WC) and/or waist-hip-ratio (WHR) with the latter also considered a measure of body fat distribution, are additional risk factors for cardiovascular disease providing information independent from BMI^7,9^.

Recent meta-analyses of genome-wide association studies (GWASs) identified hundreds of significant risk loci for BMI, WC and WHR^10,11^. The single-nucleotide polymorphisms (SNPs)-based heritability was 0.28 for BMI^10^, 0.12 for WC^11^, and 0.19 for WHR^10^, amounting to one third to half of the heritabilities estimated in family or twin studies^2,11,12^. Findings of GWASs are often used to calculate polygenic risk scores (PRSs). The PRS is a sum score of SNPs each weighted by the strength of its association with the outcome of interest as found in the GWAS^13^. A PRS is used to estimate the degree to which an individual is at risk of a disease due to genetic makeup^13^. However, because the PRS captures only part of the genetic contribution explained by common genetic variants, imperfect prediction of outcomes, like BMI, WC or WHR, is to be expected^13^. Even if all common SNPs for a particular outcome would have been identified (i.e., in case of a saturated map of common genetic variants such as available for body height^14^), the presence of additional risk factors (e.g., dietary habits or physical activities) inevitably contribute to diseases.

In the pre-GWAS era, researchers often used family history to predict an outcome. For example, offspring showed greater increases in BMI if parents had higher BMI^15–17^. In addition, based on a multigenerational family design, we found that individuals with obese first-degree relatives have 1.88 times higher risk to become obese themselves^12^. Family history captures additional genetic effects beyond those from common variants (SNPs) in the PRS as well as shared environmental influences, such as dietary habits and physical activities^5^. Family history may thus be a more potent predictor, yet the PRS has potentially less measurement error^13^. It follows that when both the PRS and family history are used, we may improve the prediction of (indices of) obesity. In addition, effects of family history including shared family environment may moderate genetic effects on obesity. Therefore, the effect of the interaction between family history and PRSs on indices of obesity is also estimated in the current study.

Maternal obesity has consistently demonstrated a correlation with an increased risk of obesity (or higher BMI) in both childhood and adulthood among offspring^16,17^. Notably, a recent study found that an elevated maternal PRS for BMI was associated with higher birth weight in offspring^18^. This effect of PRS reflects direct genetic transmission, as each parents transmits half of their genetics to their offspring during meiosis. Moreover, research has indicated that alleles not transmitted to offspring during meiosis, which are encompassed within the maternal and paternal PRSs, may still influence offspring indirectly through their impact on parental behaviors^19^. Consequently, parents shape the familial environment consistent with their genotypes, subsequently influencing the development of their offspring^20^. Although this phenomenon has been observed for educational attainment^19^, the extent to which indirect genetic effects from parents to offspring can be identified regarding indices of obesity remain uncertain. The present study investigated the relative effects of parental PRS (via offspring’s PRS) and parental obesity phenotypes (i.e., BMI, WC and WHR) on those of the offspring using structural equation modeling (SEM).

As mentioned, GWASs based on samples of unrelated individuals not only capture the effects of inherited genetic variations (direct genetic effects), but also indirect effects resulting from gene-environment correlation (parents shaping the environment in line with their own genotype)^21^. PRS derived from GWAS can be influenced by potential bias such as population stratification (unless adequately controlled for^22^) and by assortative mating^23^, which refers to partner selection based on similarities in certain characteristics^24^. To mitigate potential sources of bias, we can use the within-family design among siblings, accounting for the fact that siblings share the same parents and the family environment. However, it should be noted that PRS is expected to provide imperfect predictions of phenotypes. Therefore, in the within-family model we also include family history as a predictor, which may enhance the accuracy of predictions.

The aims of this extensive family study are to investigate the relative influence of PRS and family history on indices of general and abdominal obesity using between and within family approaches. More specifically, we will 1) estimate the overall effects of PRSs, family history, and their interactions on BMI, WC, and WHR; 2) investigate the relative effects of parental PRS (via offspring’s PRS) and parental BMI, WC, and WHR on the corresponding measures in the offspring; 3) compare the effects of between- and within-family PRSs, family history, and their interactions on BMI, WC, and WHR among siblings.

## METHODS

### Study sample and design

We used data from the ongoing Lifelines Cohort Study. Lifelines is a prospective population-based cohort study that recruited over 167,000 participants in the North of the Netherlands between 2006 and 2013^25^. A follow-up second assessment took place between 2014 and 2017. The follow-up third assessment is ongoing and started in 2019. Lifelines employs a broad range of investigative procedures in assessing the biomedical, socio-demographic, behavioral, physical, and psychological factors which contribute to the health and disease of the general population, with a special focus on multi-morbidity and complex genetics. Among all participants, genetic data of over 50,000 participants are available. The Lifelines Cohort Study is conducted according to the principles of the Declaration of Helsinki and in accordance with the research code of University Medical Center Groningen and is approved by its medical ethical committee. All participants signed an informed consent form.

### Measurements

At baseline, participants aged 8 years and older were invited to one of twelve Lifelines Research sites for a physical examination by a trained research nurse^25^. During this baseline visit, height without shoes was measured with a SECA 222 stadiometer and rounded to the nearest 0.5cm. Weight without shoes and heavy clothing was measured with a SECA 761 scale and rounded to the nearest 0.1kg. Waist and hip circumference was measured with a SECA 201 measuring tape and rounded to the nearest 0.5cm. During follow-up second and third assessments, height, weight, waist circumference, and hip circumference were measured by the same procedures as at baseline.

Body mass index (BMI) was calculated as weight (kg) divided by the square of height (m^2^). Waist-hip-ratio (WHR) was calculated as waist circumference (cm) divided by hip circumference (cm). According to the standard international classification of the World Health Organization, obesity was defined as BMI ≥ 30.0 for adults, and BMI z-score ≥ 2.0 for children aged 8 to 17 based on reference data from the 1997 Dutch Growth Study^26^. Likewise, abdominal obesity based on waist circumference was defined as WC ≥ 102 cm for male adults and WC ≥ 88 cm for female adults. In addition, abdominal obesity based on WHR was defined as WHR ≥ 1.02 for males and WHR ≥ 0.90 for females. For children, abdominal obesity based on WC and WHR was defined with a specific cut-off at different ages and gender^27^. For continuous traits, average BMI, WC, and WHR during baseline, second assessment, and third assessment were used as outcomes in the present study (Table S1). For dichotomous traits, participants were defined as having (abdominal) obesity if they met cut-off criteria specified above at least once during the three assessments.

Family history (FH) was defined as the number of first-degree relatives in Lifelines with (abdominal) obesity, including father, mother, children, and siblings. To access the robustness of the family history measures across different definitions, a binary family history trait (yes/no) was defined as having at least one first-degree relative in Lifelines with (abdominal) obesity.

### Genetic data

Genome-wide genotyping was available for 55,063 participants. The first subset of 17,033 participants was genotyped using the Illumina CytoSNP-12v2 array^25^. Pre-imputation quality control was performed in which samples and variants were excluded with a call rate < 95%, as well as variants with Hardy-Weinberg equilibrium (HWE) *P* < 1×10^-4^, or minor allele frequency (MAF) < 1%, and samples with a sex mismatch, deviating heterozygosity (> 4 SD from the mean) or of non-European ancestry. A total of 15,400 samples and 265,000 SNPs were available for analysis. The second subset of 38,030 participants was genotyped using the Infinium Global Screening Array® (GSA) MultiEthnic Disease Version^25^. Standard quality control was performed on both samples and markers, including removal of samples and variants with a low genotyping call rate (< 99%), variants showing deviation from HWE (*P* < 1×10^-6^) or excess of Mendelian errors in families (> 1% of the parent-offspring pairs), and samples with a sex mismatch, and very high or low heterozygosity. After quality control, a total of 36,339 samples and 571,420 SNPs were available for analysis. These two genotyping datasets were imputed using the HRC panel v1.1 at the Sanger imputation server^28^, and variants with an imputation quality score higher than 0.4 for variants with a MAF > 0.01 were retained. After removing duplicate samples between the two genetic datasets (n=937), 50,802 participants with genetic data were available (Supplementary Figure S1).

### Polygenic risk scores

PRSs were calculated using the genome-wide genetic data of the Lifelines participants and summary statistics of recent large GWAS meta-analyses for BMI^10^, WC^29^, and WHR^10^ in PLINK v1.9^30^ and R 4.0.3^31^ (Table S2). PLINK removed strand-ambiguous SNPs and pruned our target sample to obtain independent SNPs using clumping (*r*^2^=0.1, within a 1000 kb window). Independent risk alleles in dosage were weighted by the allelic effect sizes estimated in the summary statistics and aggregated into PRSs in R 4.0.3. PRSs were generated for eleven *P* thresholds: < 5×10^-8^, < 1×10^-7^, < 1×10^-6^, < 1×10^-5^, < 1×10^-4^, < 0.001, < 0.01, < 0.05, < 0.1, < 0.5, ≤ 1.0, determined by the summary statistics and standardized. Further, we performed principal component analysis (PCA) on the total set of 11 PRSs and used the first PRS-PC in all analyses^32^.

### Analytical approach

To address our first aim, we estimated the effects of PRSs, family history and their interactions on BMI, WC, and WHR using linear mixed regression models. Second, to investigate the relative effect of parental PRS (both via offspring’s PRS and directly) and parental obesity phenotypes (BMI, WC, and WHR) on the corresponding measures in offspring, we used SEM to construct parent-offspring transmission paths. Third, to investigate potential biases arising from indirect genetic effects, such as population stratification and assortative mating, we used linear mixed regression models to estimate the effects of between- and within-family PRSs, family history, and their interactions on BMI, WC and WHR.

### Statistical analyses

To access the relationship between increased genetic factors and the patterns observed in prevalence and family history, we calculated the prevalence of (abdominal) obesity and percentages of participants with a binary family history (yes/no) of (abdominal) obesity across different PRS deciles. For continuous traits, linear mixed regression models were used to estimate the additive effects of family history and PRS, as well as their interaction on respectively BMI, WC, and WHR. Logistic regressions were used for binary traits, specifically to estimate the effect of PRS (top vs bottom deciles) in combination with binary family history on (abdominal) obesity. Additionally, to check the genetic overlap between PRS and family history, we run linear regression of PRS on the family history (number of first-degree relatives) to estimate the portion of variance in family history that can be explained by PRS.

SEM was used to estimate the path coefficients depicting the transmission from parents to offspring (Figure 1). Paths *ma* and *pa* represent the effects of parental PRS on parental phenotypes. Paths *mb* and *pb* represent the effects of parental PRS on offspring’s PRS. Path *c* represents the effect of offspring’s PRS on offspring’s phenotype. Paths *md* and *pd* represent the effects of parental phenotype on offspring’s phenotype. Paths *me* and *pe* represent the interactions of parental phenotypes (PP) and offspring’s PRS on offspring’s phenotype. The primary analyses were conducted among participants with both parents in Lifelines (n=2,003), where paternal and maternal effects and phenotypic and PRS partner correlations were estimated in one SEM model. Residualized PRSs adjusted for chips (GSA vs CytoSNP) and 10 principal components, and standardized phenotypes were used in the SEM model. SEM was clustered using Family ID. Paternal and maternal effects were estimated separately in the larger sized samples of 5,343 father-offspring pairs and 6,920 mother-offspring pairs to evaluate robustness of findings. To investigate whether potential partner similarities were due to assortative mating, additional phenotypic and PRS correlations were estimated among 4,894 partner pairs in Lifelines (Supplementary materials).

**Figure 1.**
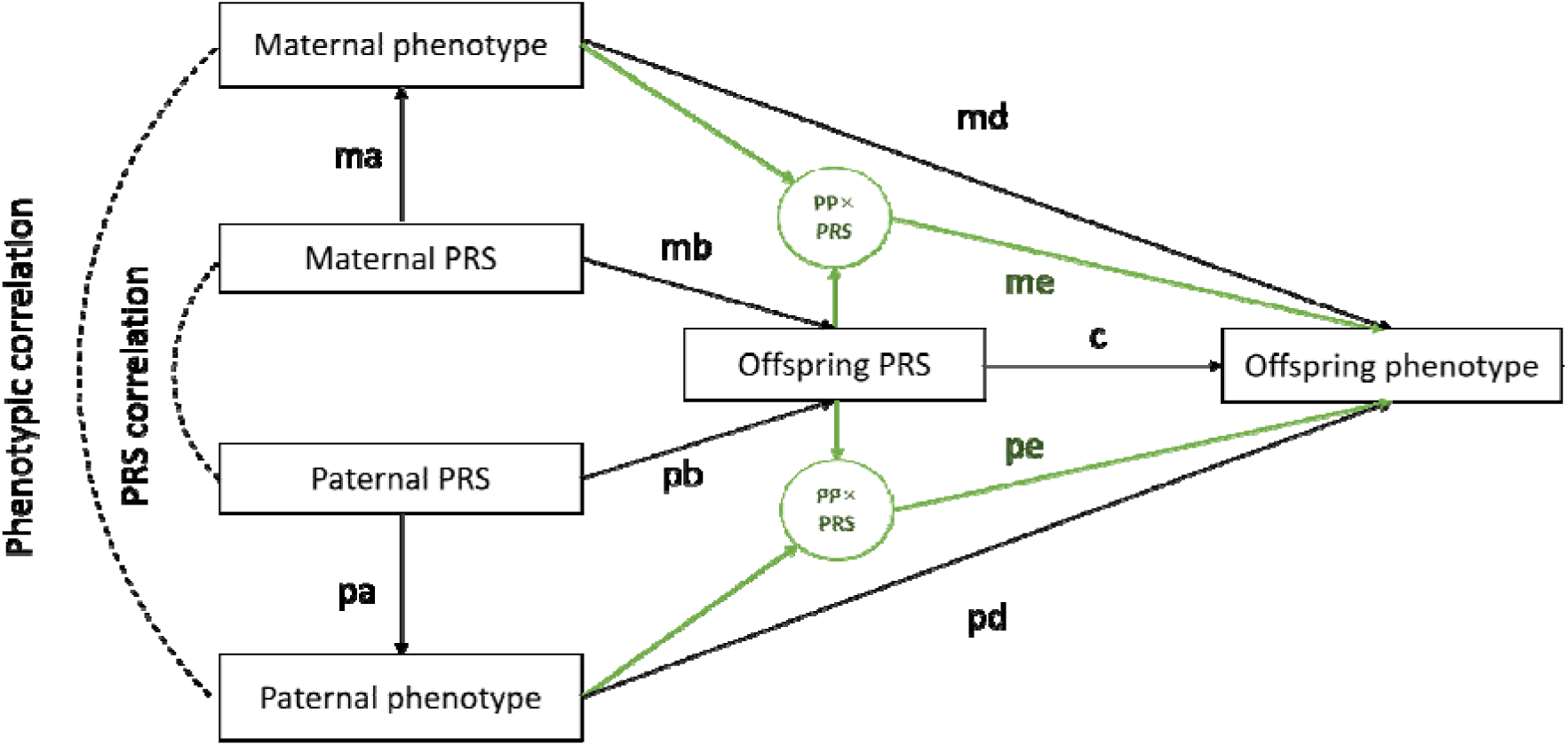
SEM parent to offspring transmission model. PP, parental phenotype; PRS, polygenic risk score.

In the Lifelines Cohort Study, a total of 17,680 participants who had at least one sibling were genotyped. In case where participants had multiple siblings, we randomly selected two siblings, resulting in a final sample size of 7,916 sibling pairs (equivalent to 15,832 participants) for our sibling analyses. We applied a random intercept linear mixed model, including the two fixed effects to separate the total effect of PRSs into within- and between-sibling effects as follows^20^:

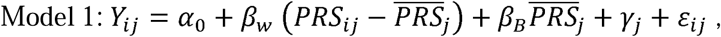

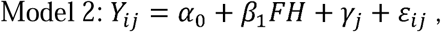

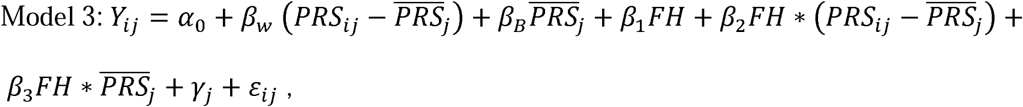

where Y is the outcome, PRS is the polygenic risk score, FH is the family history, *i*= {1,2} depicts the two siblings *i* clustering within family *j*, and 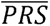 refers to the mean of PRS value in family *j*. The α_0_ represents the intercept and *γ_i_* the random effect of family *j*, and ε_*ij*_ the residual for each sibling *i* in family *j*. The between-siblings effect *β_B_* represents the expected change in the outcome Y given a one-unit change in the average of the family PRS, and the within-siblings effect *β_w_* represents the expected change given a one-unit change in the difference between the individual PRS and the average PRS of the siblings. *Β_1_* represents the main effect of family history on the outcome. *Β_2_* and *β_3_* represent the interaction effects between family history and respectively within- and between-family PRSs. We performed the same analyses among all siblings (n=17,680) to check consistency of effects.

For all analyses, a p < 0.05 based on two-sided testing was considered statistically significant. All analyses were conducted in R4.0.3.

## RESULTS

Among all participants of Lifelines from European ancestry, 50,747 participants provided information on both genetic data and measurement of weight, height, waist circumference, and hip circumference (Figure S1). Table 1 shows characteristics of the participants. They had a mean age of around 45 years and 58% were female. The prevalence of general obesity was 18.04%. Prevalences of abdominal obesity were 40.72% and 39.04% based on WC and WHR, respectively.

**Table 1.**
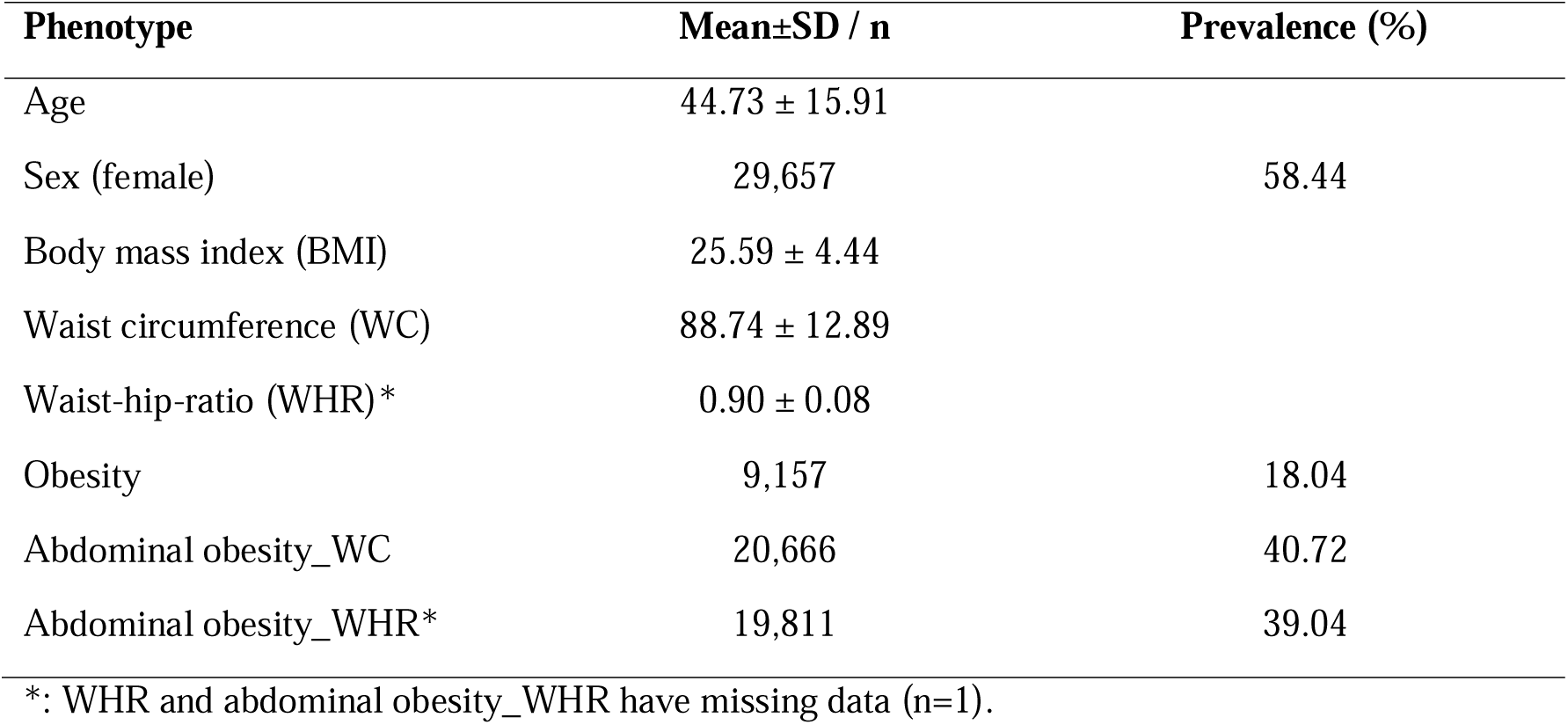
Characteristics of participants (n=50,747)

Polygenic risk scores (PRSs) explained 8.75%, 5.19% and 2.92% of the variance in BMI, WC and WHR, respectively, which were larger than those explained by family history (4.28% for BMI, 2.69% for WC, and 0.89% for WHR, Figure 2). When combined into one model, PRS and family history together explained 11.81%, 7.26% and 3.62% of the variance in BMI, WC and WHR, respectively (Figure 2). In addition, significant interactions between family history and PRSs were found for BMI (*p*=5.27×10^-8^) and WC (*p*=5.40×10^-4^), but not for WHR (*p*=0.21). These interactions explained little additional variance (Figure 2). Similar results albeit with lower variance explained by family history was found if family history was expressed as a binary (yes/no) variable (Figure S2). Small but statistically significant genetic overlap was observed between PRS and family history defined as the number of first-degree relatives with (abdominal) obesity with PRS explaining 2.33% of the variance in the family history of general obesity, and 1.42% and 0.76% in the family history of abdominal obesity based on WC and WHR, respectively (Figure S3). With increased PRS deciles, higher prevalences of (abdominal) obesity and higher percentages of participants with family history were found (e.g., from the bottom to the top PRS decile, the prevalence of obesity increased from 5.36% to 37.36%, and the percentage of participants with family history (yes/no) increased from 17.94% to 41.48%, Figures S4 and S5). Compared with individuals with a PRS in the bottom decile, individuals with a BMI PRS in the top decile had 2.45 times higher risk to become obese, and the risk increased to 3.28 times if individuals additionally had at least one first-degree relative with obesity (Figure S6).

**Figure 2.**
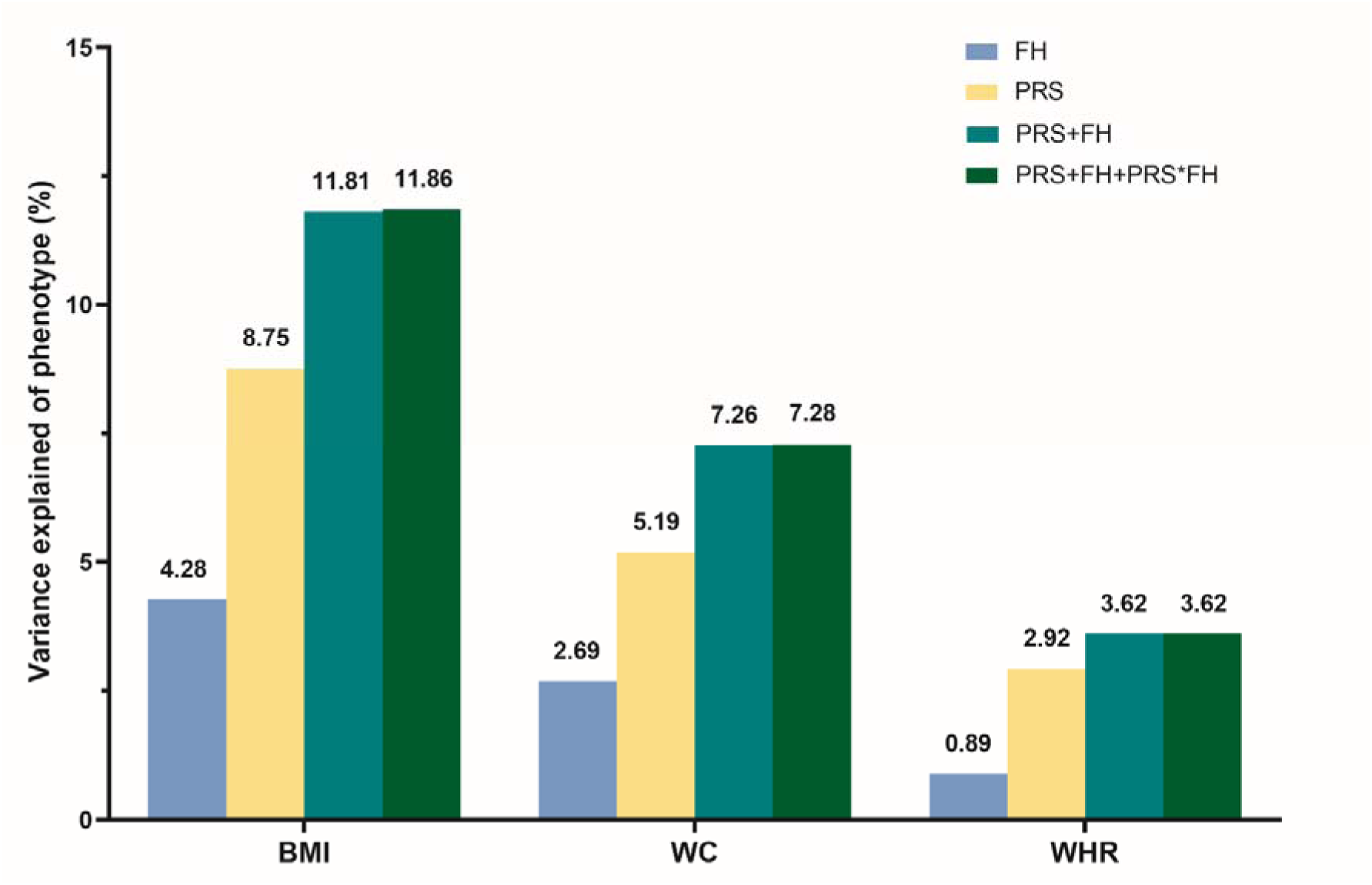
The variance explained by FH and PRS for BMI, WC, and WHR. FH, family history, the number of obese first-degree relatives; PRS, polygenic risk score; BMI, body mass index; WC, waist circumference; WHR, waist-hip-ratio. The effects of family history, PRS and their interaction were significant for BMI, WC and WHR (p<0.05), except for the interaction between PRS and family history for WHR.

For parent-offspring transmission, among 2,003 participants with both parents participating in Lifelines, the expected halves of maternal and paternal PRSs transmitted to offspring’s PRS (*β*=0.48-0.52, paths *mb* and *pb*, Figure 3). Offspring’s PRS, in turn, had a significant effect offspring’s BMI, WC and WHR (*β*=0.14-0.16, path *c*, Figure 3). Independent of this parent-offspring transmission pathway based on the PRS, high maternal and paternal BMI, WC and WHR were significantly associated with high offspring’s BMI, WC and WHR (*β*=0.14-0.27, paths *md* and *pd*, Figure 3). Effects sizes were similar for maternal and paternal effects of BMI, WC and WHR on the corresponding obesity indices in offspring. Interactions between parental phenotypes and PRSs of offspring were non-significant (paths *me* and *pe*, Figure 3). Phenotypic correlations between parents were 0.23 for BMI, 0.27 for WC and 0.16 for WHR. PRSs correlations between parents were 0.07 for BMI, 0.06 for WC and 0.01 for WHR. When additional analyses were performed separately for 6,920 mother-offspring pairs and 5,343 father-offspring pairs, similar parent-offspring transmission results were found (Figure S7). In addition, similar phenotypic and PRS correlations were found among 4,894 partner pairs in Lifelines, with phenotypic correlations of 0.26, 0.31 and 0.23 and PRS correlations of 0.04, 0.03, 0.01 for BMI, WC and WHR, respectively (Figure S8-9).

**Figure 3.**
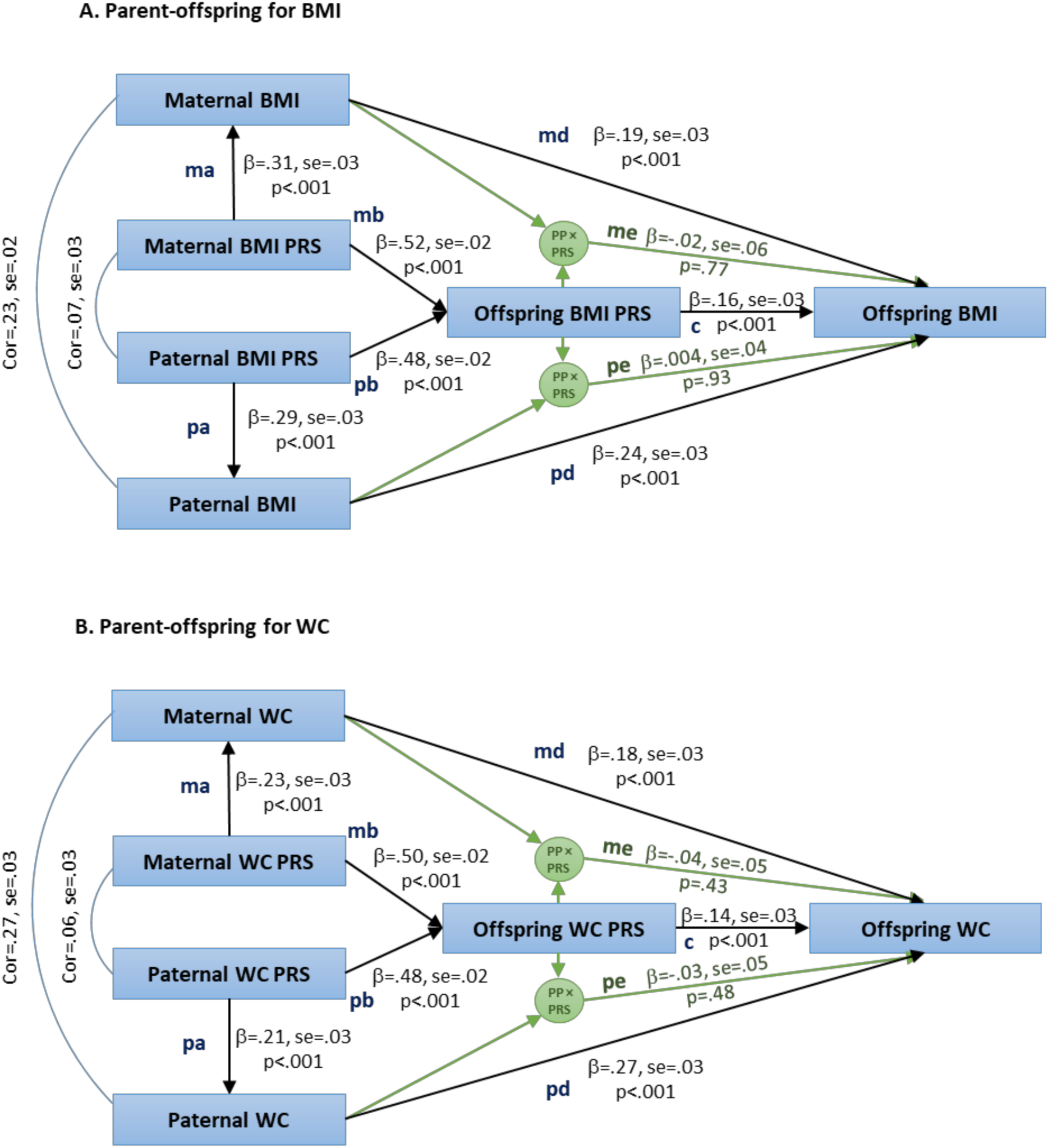
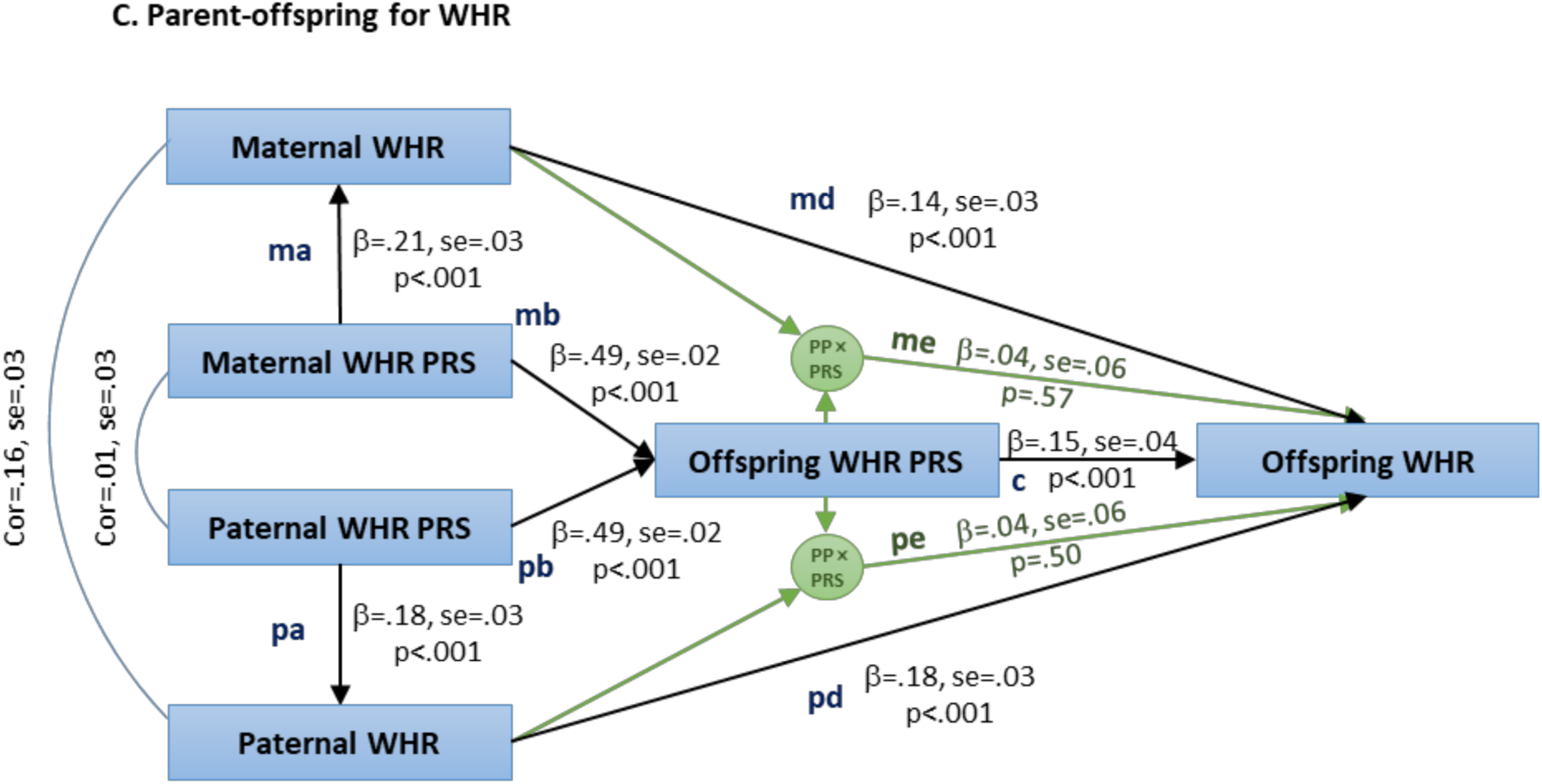
Parent-offspring transmission path of BMI, WC, and WHR among participants with both father and mother in Lifelines. PP, parental phenotype; PRS, polygenic risk score; BMI, body mass index; WC, waist circumference; WHR, waist-hip-ratio. There were 2,003 parents-offspring pairs for BMI, WC, and WHR. PRS was adjusted for chips (GSA vs CytoSNP), and 10 principal components. SEM was clustered using Family ID.

To test between- and within-family PRS effects, we included 7,916 sibling pairs participating in Lifelines. Between-family PRSs had similar effect sizes as within-family PRSs (i.e., *β_B_*=0.30 and *β_W_*=0.28 for BMI, *β_B_*=*β_W_*=0.23 for WC, and *β_B_*=0.19 and *β_W_*=0.17 for WHR, model 1, Figure 4, Table S3). Between-family PRSs explained more variance for BMI, WC, and WHR than within-family PRSs (i.e., 6.70% vs 1.91% for BMI, 4.10% vs 1.25% for WC, and 2.61% vs 0.72% for WHR, model 1, Figure 4, Table S3) which is according to expectation (detailed formula in supplementary). In model 2, family history explained 5.07% for BMI, 3.12% for WC, and 1.32% for WHR. In model 3, the combination of family history and between- and within-family PRS and their interactions explained more variance than individual predictors, with total variances explained of 11.72% for BMI, 7.52% for WC, and 4.26% for WHR. In addition, for BMI and WC, interactions between family history and between- and within-family PRSs were significant but had small effect sizes (*β*=0.02-0.06, model 3, Figure 4). Similar results were found in the sensitivity analyses among all siblings (Table S4).

**Figure 4.**
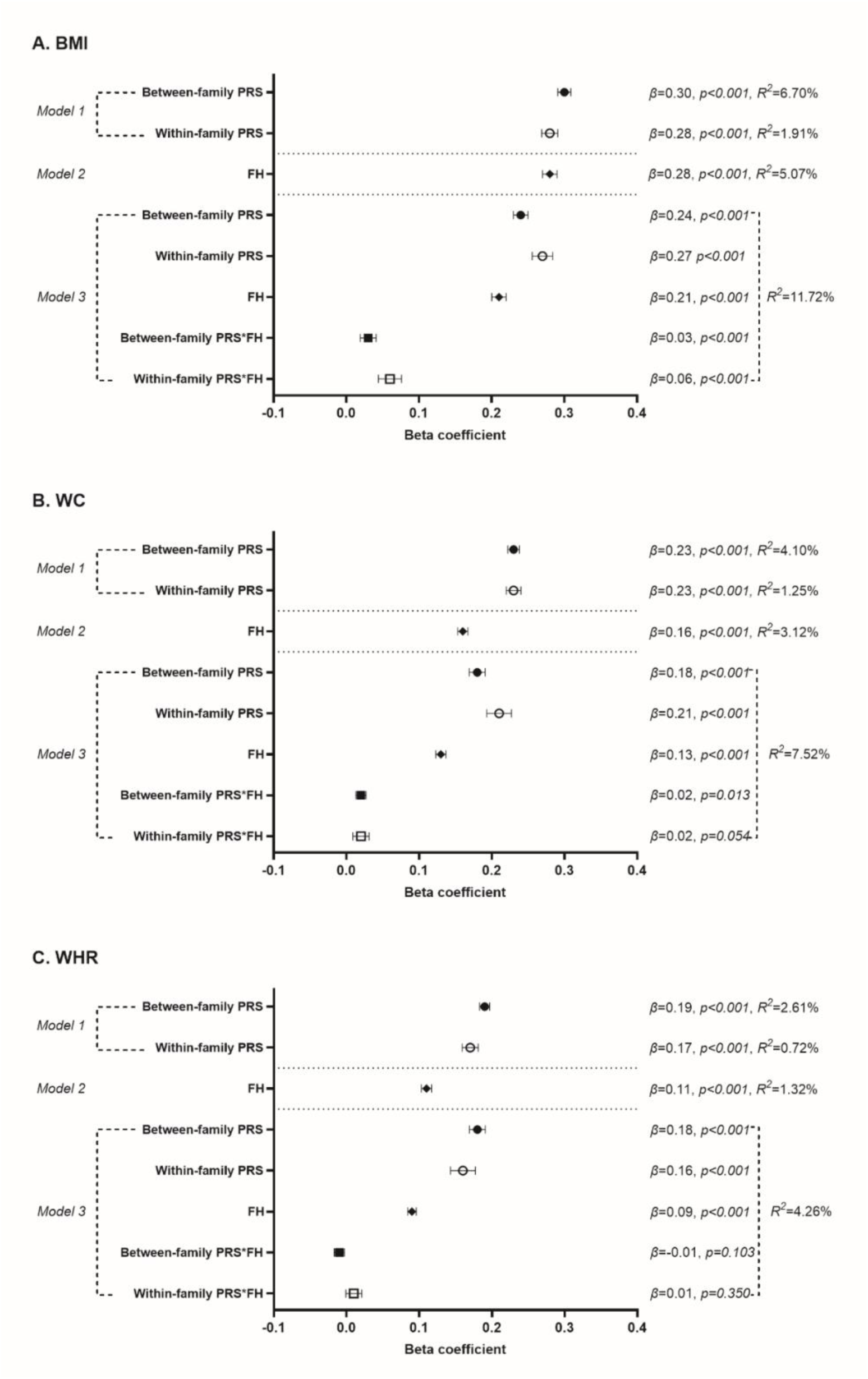
The effect of between- and within-family PRSs and family history on BMI, WC, and WHR among 7,916 sibling pairs. FH, family history; PRS, polygenic risk score; BMI, body mass index; WC, waist circumference; WHR, waist-hip-ratio. We adjusted age, sex, chips (GSA vs CytoSNP) and 10 principal components in model 0. *R^2^* represented the variance explained by PRS, or FH, or their combination in addition to model 0. The variance explained by between- and within-family PRS in line with the theory (detail in supplementary).

## DISCUSSION

In this large family study, we found that PRS and family history together explained more variance of obesity indices BMI, WC and WHR than a single predictor. In addition to transmission of the PRS from parent to offspring, high parental BMI, WC and WHR were independently associated with high offspring’s BMI, WC and WHR. Between-family PRSs had similar effect sizes as within-family PRSs for BMI, WC and WHR, which indicated that potential biases on the genetic effects of these obesity indices such as caused by population stratification, assortative mating and shared family environment were very small.

It is well known that parental obesity (or higher BMI) is associated with offspring obesity or overweight^16,17,33^. However, few studies have considered both genetic transmission and parental phenotypic effects in one study^34^, as most studies do not have genetic data for both parents and offspring. Half of the paternal and maternal PRSs were transmitted to offspring’s PRS and significantly influenced offspring’s obesity indices, which was consistent with previous findings that paternal and maternal transmitted genetic alleles influence offspring BMI or overweight equally^19,34^. The combination of PRS and family history improved the prediction of general and abdominal obesity. Consistent with previous studies^35^, PRSs only explained a relatively small part of the total genetic effects on these obesity indices, i.e., they explained less than half of the known SNP heritabilities^10, 11^ and less than one-fifth of the pedigree heritabilities^12^. Further, we found that independent of the PRS transmission from parent to offspring, high parental BMI, WC and WHR were associated with high BMI, WC and WHR among offspring. The reason is that PRSs only capture part of the effect of common genetic variants identified in GWAS, while family history likely reflects additional genetic effects not captured by PRS (i.e., remaining common genetic variants and rare variants^36^ as well as effects of the shared family environment. In line with this, the overlap between them was small, with PRS explaining only 2.33% of the family history of obesity. With the quality and sample size of GWASs steadily improving, the variance explained by PRS will also increase and approach the SNP-based heritability^13, 14^. In addition, Wainschtein et al^36^ found that including rare variants from whole-genome sequence data (i.e., MAF>0.0001), the SNP-based h^2^ increased from 0.24 based on common variants to 0.29 for BMI, where the rare variants increased 17% of SNP-based h^2^. With the application of whole-genome sequence data, we may will identify the remaining common genetic variants and rare variants as currently captured by family history.

Individuals with higher PRSs had higher prevalence of (abdominal) obesity as well as a higher likelihood of a positive family history, compared to those with lower PRSs. However, over half of the participants in the top BMI PRS decile were not obese (62.64%) and had no family history of obesity (58.52%). This indicates that even with a high genetic risk (i.e., the upper 10% of the PRS), an individual may not have obesity or have no family history. Obviously genetic and familial factors are not the only contributors to risk of obesity, they act in conjunction with environmental factors, such as individual dietary habits and physical activity^4,5^. Furthermore, due to the segregation of genetic variants at meiosis PRSs vary between family members^13^. According to the polygenic theory, most people with common disease have no known affected family members due to the reconciliation of both the polygenic nature of common disease and the frequency of a disease in the population^37^. Therefore, individuals with high BMI PRSs, but without obesity or a family history, are expected.

Although most research has focused on the role of maternal obesity on offspring obesity^16–18,36^, the present study found that paternal and maternal obesity phenotypes influence offspring obesity phenotypes equally. Consistent results were found in a Finnish study (1,788 child-mother-father trios), where paternal and maternal pre-pregnancy obesity had a similar effect on offspring overweight at the age of 16 years (father-offspring OR=3.17-5.58; mother-offspring OR=3.95-4.36)^33^, as well as a study from Brazil in which maternal and paternal BMI were equally associated with their adult offspring obestatin levels (i.e. obestatin is a biomarker in regulation of appetite and energy homeostasis, glucose, and lipid metabolism)^38^. However, a UK study found that maternal BMI had a stronger correlation with the BMI of adult daughters in comparison to paternal BMI, while both maternal and paternal BMI had equivalent associations with the BMI of adult sons^15^. According to the “fetal overnutrition hypothesis”, maternal obesity is likely to be a stronger influencing factor due to the intrauterine environment^39^. In relation to the hypothesis, maternal obesity effects may have stronger influence on infant and early childhood BMI^40^, while this influence may be less further on in development given that most studies with family trio designs showed similar effect sizes of maternal and paternal obesity on for adolescent and adult offspring obesity^15, 33, 38^.

Between-family PRSs had similar effect sizes as within-family PRSs for BMI, WC and WHR among siblings. Similar effect sizes of between and within-family BMI genetic risk scores were also found for the BMI differences among 1,353 dizygotic twin pairs (*β_B_*=0.35, 95%CI=0.31-0.39; *β_W_*=0.30, 95%CI=0.24-0.36)^20^. In addition, a recent within-sibship GWAS (n=178,076) found that within-sibship SNP heritability for BMI attenuated by only 4.76% from the population estimate (population h^2^=0.21, 95%CI=0.19-0.23; within-sibship h^2^=0.20, 95%CI=0.17-0.23; difference P=0.27)^21^. Consistent results were also shown across different populations. In an East Asian population (n=13,856), the percentage shrinkage in effect size of the within-sibship model for BMI was less than 20% compared with the population model^21^. Furthermore, Kong et al found that parental non-transmitted PRS of BMI-associated alleles was not associated with offspring BMI^19^, and similar results were also found in a Danish study where the maternal non-transmitted genetic risk score was not associated with child overweight (OR=0.98, 95%CI=0.88-1.10)^34^. In line with the non-significant effect of non-transmitted alleles from parent to offspring BMI or overweight, the consistently similar effect sizes of between and within-family PRS in siblings indicated negligible bias for the genetic effect for indices of obesity such as caused by population stratification, assortative mating and shared family environment.

The following limitations of our study need to be considered. First, not all first-degree relatives of Lifelines participants took part in the study, which may have underestimated the effect of family history. Second, this study was conducted among Europeans, which is difficult to generalize to other ancestries. The present study also has a few numbers of strengths. First, we defined the family history based on the accurate measurements of obesity indices for all participants in Lifelines, which is more precise than self-reported family history. Second, the Lifelines Cohort Study has a large sample size, multi-generational family design, and genome-wide data of many participants, which offered the opportunity to investigate the relative influence of PRS and family history on general and abdominal obesity using between and within-family approaches.

In summary, the present study finds that the combination of PRS and family history improved the prediction of indices for general and abdominal obesity. In addition to the genetic effect captured by the parent-offspring PRS transmission, parental BMI, WC and WHR also independently influenced offspring’s BMI, WC and WHR, which may reflect additional genetic effects not captured by PRS as well as effects of the shared family environment. Similar effect of between- and within-family PRS between siblings offers the evidence that negligible bias for genetic effect for indices of obesity such as caused by population stratification, assortative mating, and shared family environment. Our findings are relevant for obesity risk screening in the clinical context. Given that the PRS can potentially be estimated early in life, implementing preventive measures at an early stage may be helpful, for instance, if there is a high risk of obesity among family members^41,42^, although this approach also raises ethical considerations and potential for (self-)stigmatization^43^. Nonetheless, more generally we believe that our findings on individual differences in genetic and familial risk are part of an emerging literature that may contribute to the reduction of societal stigma associated with obesity.

## Data Availability

All data produced in the present study are available upon reasonable request to the authors

https://wiki.lifelines.nl/doku.php

## ACKNOWLEDGEMENTS

We acknowledge the services of the Lifelines Cohort Study, the contributing research centres delivering data to Lifelines, and all the study participants.

## AUTHOR CONTRIBUTIONS

RW, CAH, PMV, HS contributed to the study conception and design. Lifelines Cohort Study offered the data. RW did data analysis and drafted the manuscript. All authors participated in revising it critically for important intellectual content.

## FUNDING

The Lifelines Biobank initiative has been made possible by funding from the Dutch Ministry of Health, Welfare and Sport, the Dutch Ministry of Economic Affairs, the University Medical Center Groningen (UMCG the Netherlands), University of Groningen and the Northern Provinces of the Netherlands. The generation and management of GWAS genotype data for the Lifelines Cohort Study is supported by the UMCG Genetics Lifelines Initiative (UGLI). UGLI is partly supported by a Spinoza Grant from NWO, awarded to Cisca Wijmenga. RW acknowledges support from the China Scholarship Council (201806010404).

## COMPETING INTERESTS

The authors declare no competing interests.

## Notes

### Competing Interest Statement

The authors have declared no competing interest.

### Author Declarations

The Lifelines Cohort Study is conducted according to the principles of the Declaration of Helsinki and in accordance with the research code of University Medical Center Groningen and is approved by its medical ethical committee.

