## Supplementary for "The effect of family history and polygenic risk scores on general and abdominal obesity in the Lifelines Cohort Study"

[3]A full list of members and affiliations appears in the Supplementary Note

[4]Institute for Molecular Bioscience, The University of Queensland, Brisbane, Queensland, Australia

[5]Big Data Institute, Li Ka Shing Centre for Health Information and Discovery, Nuffield Department of Population Health, University of Oxford, Oxford, UK.

[*]Corresponding author: University of Groningen, University Medical Center Groningen, Department of Epidemiology, P.O.Box 30.001 (FA40) Hanzeplein 1, 9700 RB Groningen, Netherlands. Phone: +31657335908. E-mail address:

### Supplementary Note

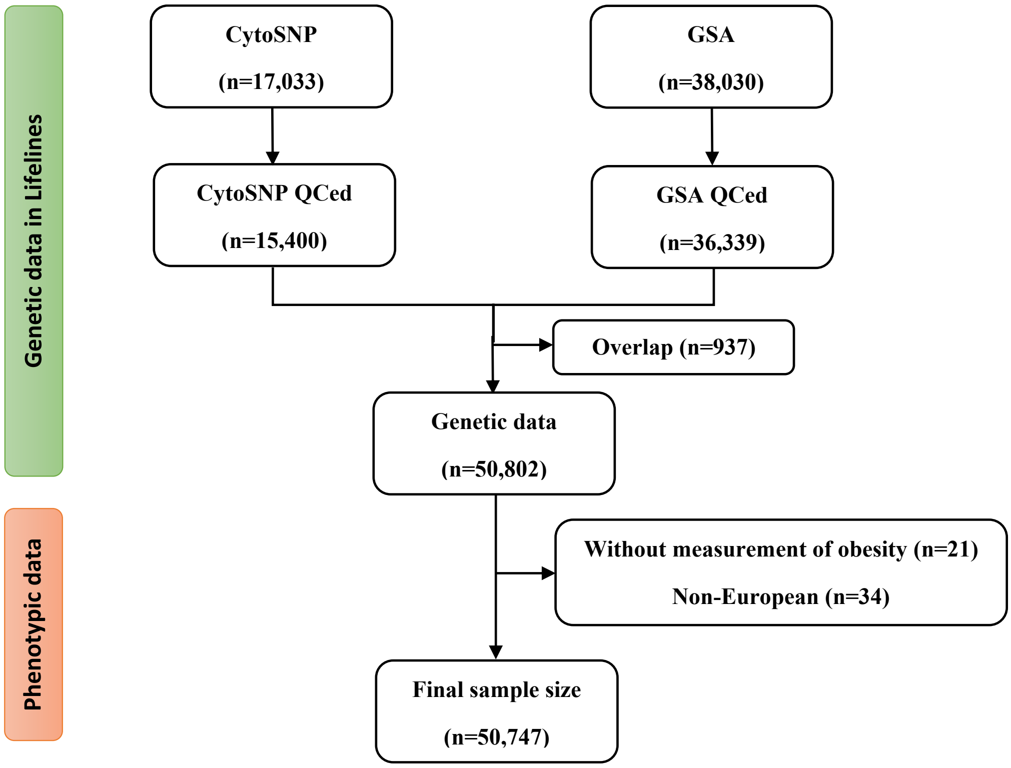


**Figure.S1 Flow chart** CytoSNP, Illumina CytoSNP-12v2 array; GSA, Infinium Global Screening Array® (GSA) MultiEthnic Disease Version.

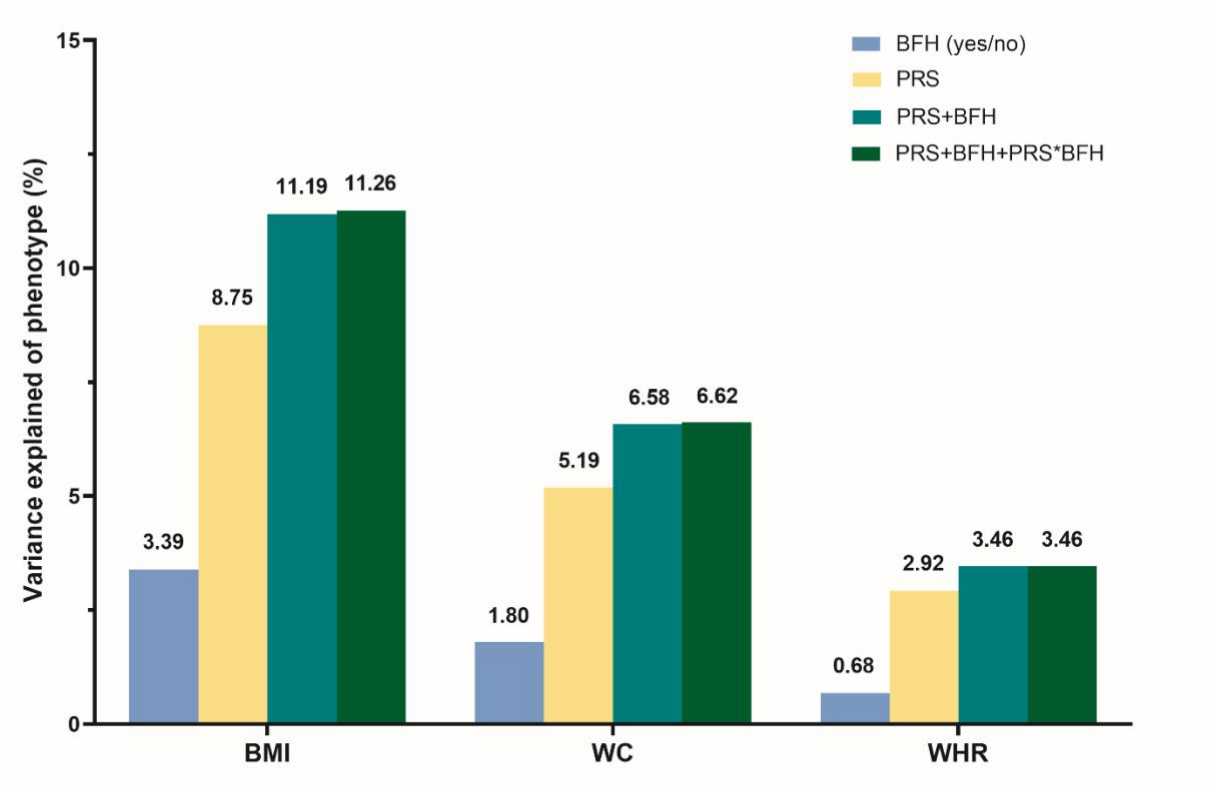


**Figure S2. The variance explained by binary family history and PRS for BMI, WC, and WHR.** BFH, binary family history (yes/no); PRS, polygenic risk score; BMI, body mass index; WC, waist circumference; WHR, waist-hip-ratio. The effects of binary family history, PRS and their interaction were significant for BMI, WC and WHR ($p<0.05$), expect for the interaction between PRS and binary family history for WHR.

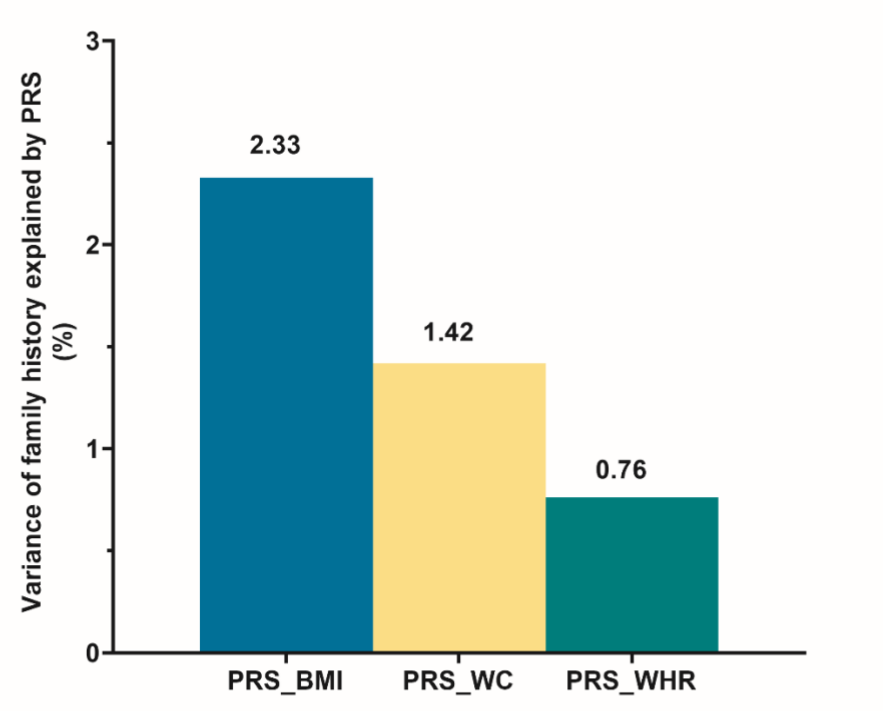


**Figure S3. Variance explained in family history defined as the number of first-degree relatives in Lifelines with (abdominal) obesity by PRS.** PRS, polygenic risk score; BMI, body mass index; WC, waist circumference; WHR, waist-hip-ratio.

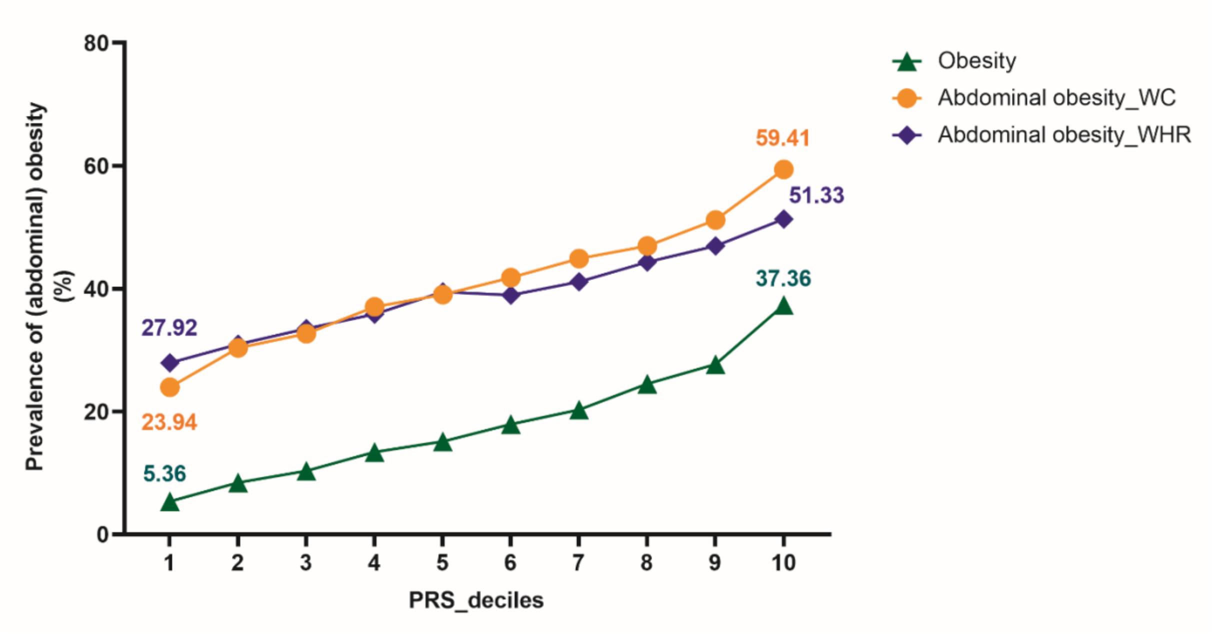


**Figure S4. The prevalence of (abdominal) obesity among different PRS deciles.** PRS, polygenic risk score; Abdominal obesity_WC, abdominal obesity defined based on waist circumference; Abdominal obesity_WHR, abdominal obesity defined based on waist-hip-ratio.

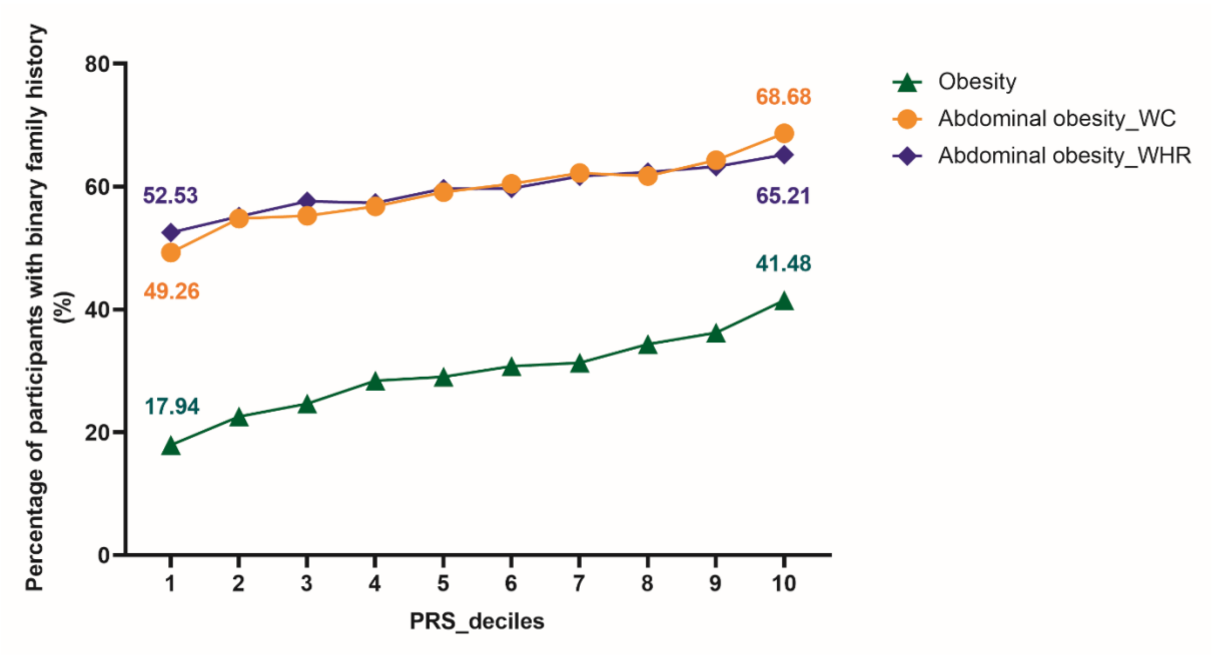


**Figure S5. The percentage of participants with binary family history at different PRS deciles.** PRS, polygenic risk score; Abdominal obesity_WC, abdominal obesity defined based on waist circumference; Abdominal obesity_WHR, abdominal obesity defined based on waist-hip-ratio.

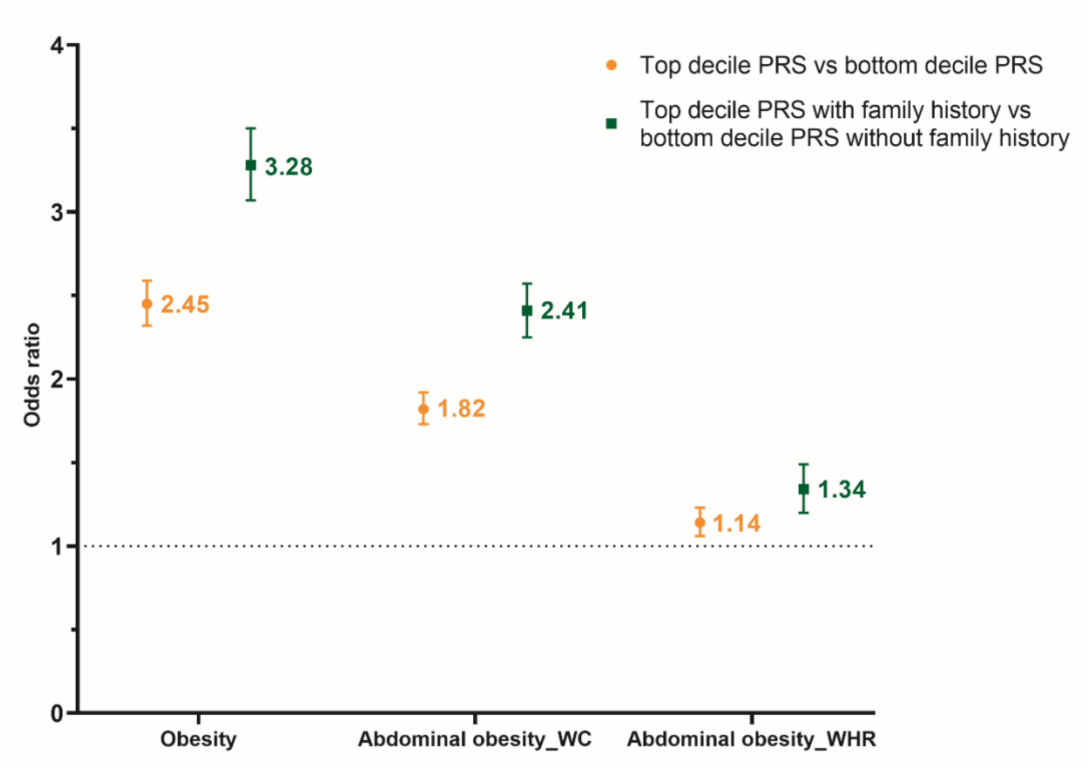


**Figure S6. The odds ratio of top versus bottom deciles of PRS and additionally taking into account family history (yes/no) for (abdominal) obesity.** PRS, polygenic risk score; Abdominal obesity_WC, abdominal obesity defined based on waist circumference; Abdominal obesity_WHR, abdominal obesity defined based on waist-hip-ratio.

**A. Parent-offspring for BMI**

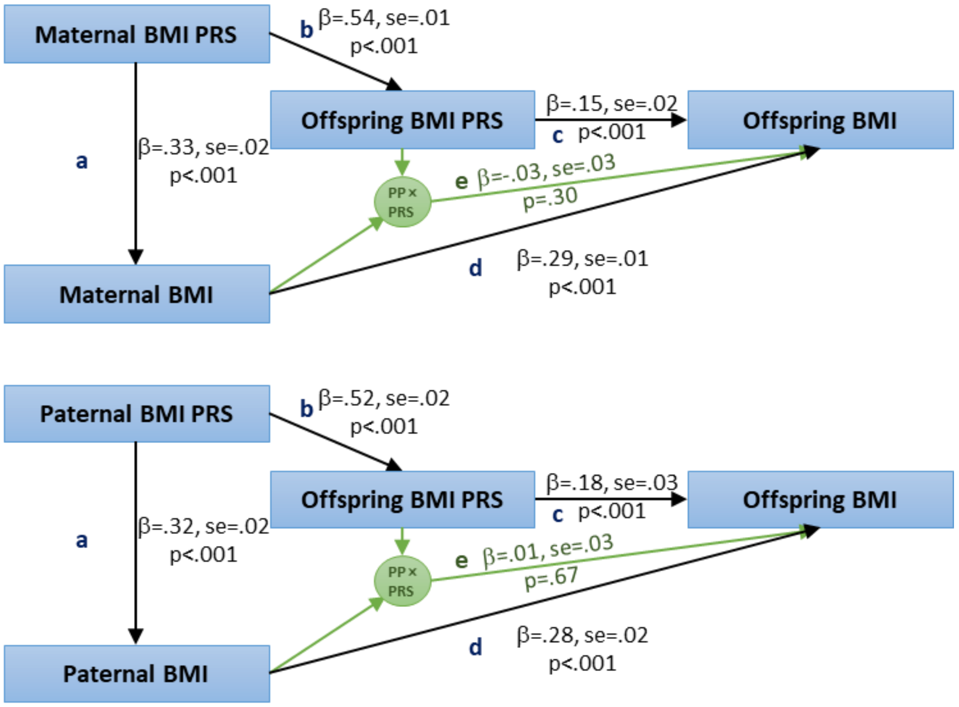


**B. Parent-offspring for WC**

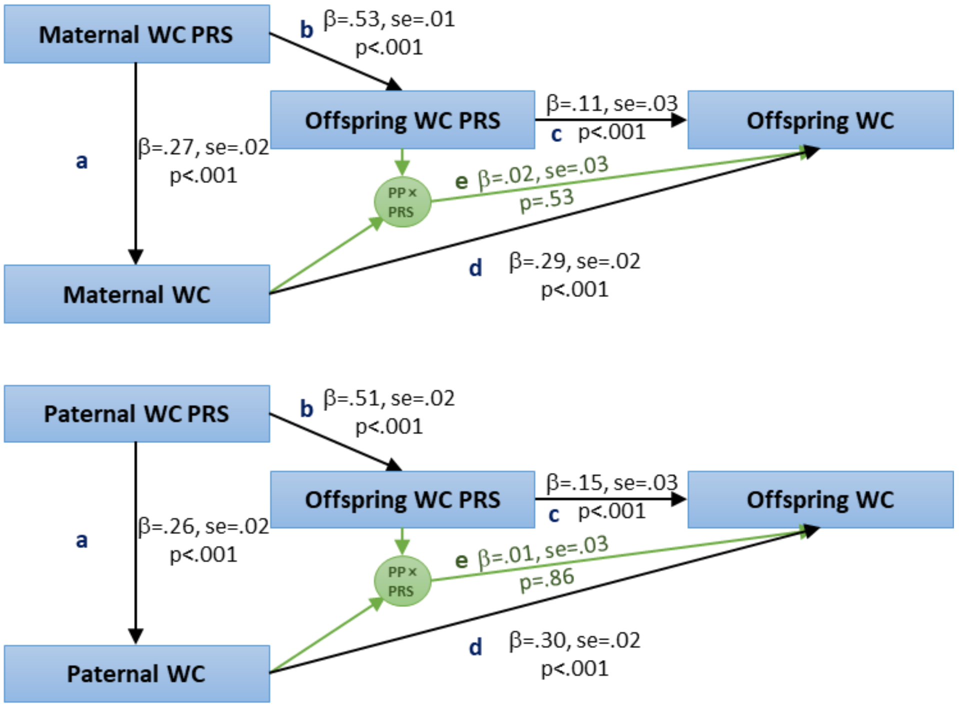

C. Parent-offspring for WHR

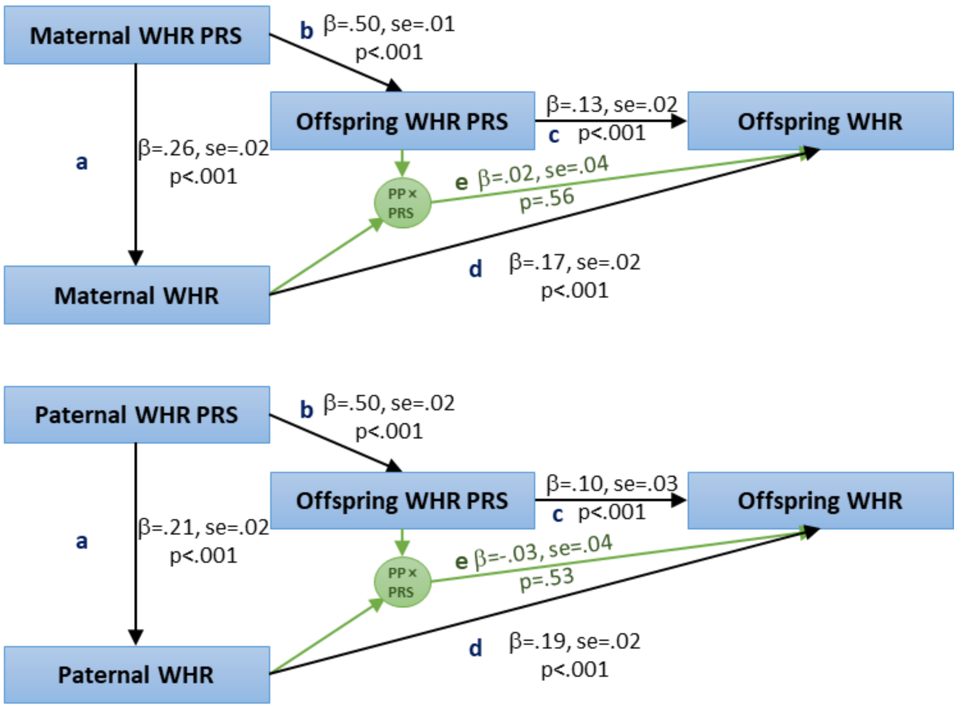


**Figure S7. Parent-offspring transmission path of BMI, WC, and WHR.** PP, parental phenotype; PRS, polygenic risk score; BMI, body mass index; WC, waist circumference; WHR, waist-hip-ratio. There were 6,920 mother-offspring pairs for BMI and WC, and 6,918 mother-offspring pairs for WHR. There were 5,343 father-offspring pairs for BMI, WC, and WHR. PRS was adjusted for chips (GSA vs CytoSNP), and 10 principal components. SEM was clustered using Family ID.

In addition to the phenotypic and PRS correlations between partners in the SEM, we also estimated the partner similarity among 4,894 pairs partners in Lifelines. Pearson correlations were used to estimate the phenotypic and PRS correlations between partners. We used $r_y$ to represent the phenotypic correlation between partners, $r_p$ and $r_m$ represent the correlations between the phenotypes and PRSs for the father and mother, respectively. The observed PRS correlation between partners should be equal to $r_y r_p r_m$ (predicted PRS correlation) if the correlation is explained by assortative mating on the phenotype alone, and the relationship between the PRS and the phenotype is linear[1]. We also estimated the correlation between the residual of the father's PRS after regression onto the father's phenotype and the residual of the mother's PRS after regression onto the mother's phenotype, which should be zero under phenotypic assortment if the relationship between the PRS and the phenotype is linear[1]. We performed further analyses adjusting for 10 principal components to test whether assortative mating on factors related to ancestry explained the PRS correlation between partners.

Among 4,894 pairs of partners, the phenotypic correlation was 0.26 for BMI, 0.31 for WC, and 0.23 for WHR (Figure S8). The observed PRS correlations between partners were quite small, and ranged from 0.01 for WHR to 0.04 for BMI (Figure S8). The predicted PRS correlations were similar to the observed PRS correlations between partners (Figure S8). In the sensitivity analysis, partner correlations between observed PRS residualized on phenotypes of partners, and partner correlations between observed PRS residualized on phenotypes and 10 principal components were nonsignificant (Figure S9).

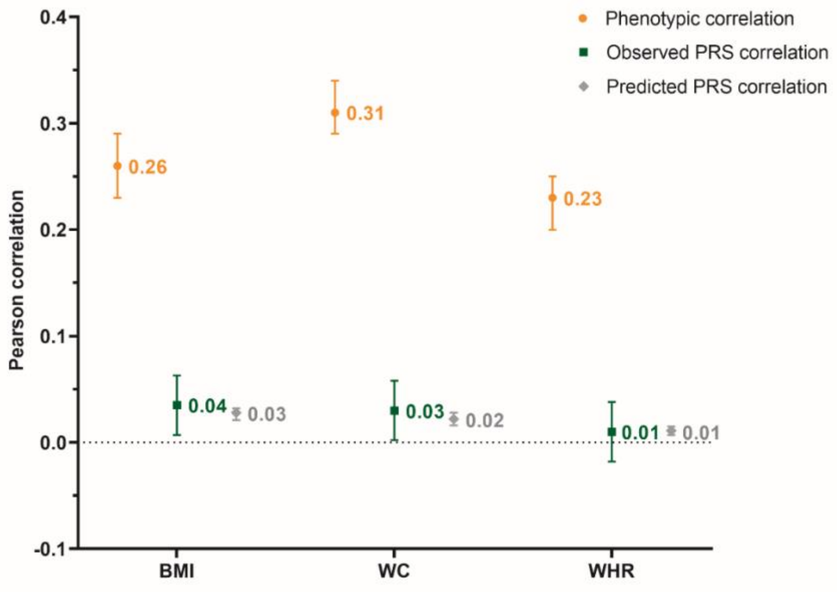


**Figure S8. Phenotypic and PRS correlations for BMI, WC, WHR between partners.** PRS, polygenic risk score; BMI, body mass index; WC, waist circumference; WHR, waist-hip-ratio. Observed PRS correltion was the direct PRS correlation between spouse. Predicted PRS correlation was defined as ry*rp*rm, where ry was the phenotypic correlation between spouse, rp and rm were the phenotype and PRS correlations for father and mother.

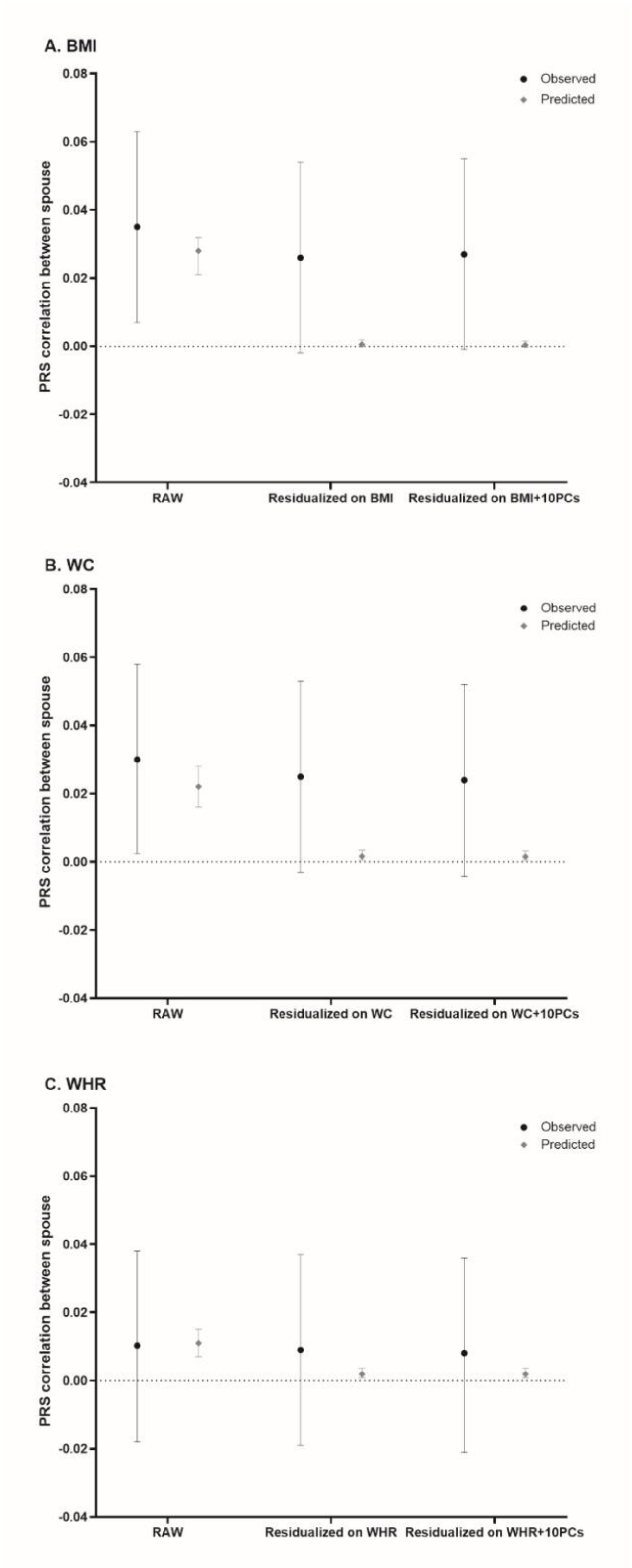


**Figure S9. PRS correlations between partners for BMI, WC, and WHR.** PRS, polygenic risk score; BMI, body mass index; WC, waist circumference; WHR, waist-hip-ratio; PCs, principal components. Observed PRS correlation was the direct PRS correlation between spouse. Predicted PRS correlation was defined as ry*rp*rm, where ry was the phenotypic correlation between spouse, rp and rm were the phenotype and PRS correlations for father and mother.

**Table S1. Means ± SD of BMI, WC, and WHR at baseline, second and third assessment**

| Phenotype | Total | | Baseline | | Second assessment | | Third assessment | |
|---|---|---|---|---|---|---|---|---|
| | n | Mean±SD | n | Mean±SD | n | Mean±SD | n | Mean±SD |
| **Age** | 50,747 | 44.73±15.91 | 50,594 | 42.23±15.42 | 41,751 | 47.18±15.32 | 16,976 | 55.56±13.23 |
| Adults | 47,627 | 46.81±14.11 | 47,206 | 44.38±13.61 | 39,668 | 48.94±13.59 | 16,832 | 55.90±12.76 |
| Children | 3,120 | 13.05±2.45 | 3,388 | 12.28±2.81 | 2,083 | 13.66±2.29 | 144 | 15.97±1.08 |
| **Gender (female)** | 50,747 | 29,658 (58.44%) | 50,594 | 29,570 (58.45%) | 41,751 | 24,429 (58.51%) | 16,976 | 10,256 (60.41%) |
| Adults | 47,627 | 28,094 (58.99%) | 47,206 | 27,822 (58.94%) | 39,668 | 23,402 (58.99%) | 16,832 | 10,183 (60.50%) |
| Children | 3,120 | 1,564 (50.13%) | 3,388 | 1,748 (51.59%) | 2,083 | 1,027 (49.30%) | 144 | 73 (50.69%) |
| **BMI** | 50,747 | 25.59±4.44 | 50,594 | 25.39±4.50 | 41,746 | 25.65±4.41 | 16,973 | 26.65±4.43 |
| Adults | 47,627 | 26.01±4.20 | 47,206 | 25.86±4.21 | 39,663 | 25.97±4.23 | 16,829 | 26.69±4.41 |
| Children | 3,120 | 19.27±3.03 | 3,388 | 18.85±3.12 | 2,083 | 19.60±3.19 | 144 | 21.59±3.72 |
| **WC** | 50,747 | 88.74±12.88 | 50,592 | 88.33±13.26 | 41,749 | 88.84±13.00 | 16,969 | 91.46±12.74 |
| Adults | 47,627 | 90.04±12.04 | 47,205 | 89.81±12.25 | 39,666 | 89.84±12.42 | 16,825 | 91.61±12.67 |
| Children | 3,120 | 68.94±8.41 | 3,387 | 67.79±9.16 | 2,083 | 69.91±8.53 | 144 | 74.80±10.12 |
| **WHR** | 50,746 | 0.90±0.08 | 50,586 | 0.90±0.08 | 41,745 | 0.90±0.09 | 16,966 | 0.90±0.09 |
| Adults | 47,626 | 0.90±0.08 | 47,199 | 0.90±0.08 | 39,662 | 0.90±0.09 | 16,822 | 0.90±0.09 |
| Children | 3,120 | 0.85±0.06 | 3,387 | 0.87±0.07 | 2,083 | 0.84±0.07 | 144 | 0.81±0.07 |

BMI, body mass index; WC, waist circumference; WHR, waist-hip-ratio.

**GWAS for obesity**

As data from some of the Lifelines subjects (n=8,118) was used in the GIANT 2015 GWAS meta-analysis for BMI and WHR[2], we used the MetaSubtract R package[3] to exclude the effect of Lifelines participants.

**Table S2. Summary statistics of recent large GWAS used in the present study**

| Outcome | GWAS | Year | Datasets | Sample size | Phenotype |
|---|---|---|---|---|---|
| BMI | Pulit et al.[4] | 2019 | UKB+GIANT-Lifelines | 798,716 | BMI |
| WHR | Pulit et al.[4] | 2019 | UKB+GIANT-Lifelines | 689,617 | WHR |
| WC | UKB[5] | 2018 | UKB | 360,564 | WC |

BMI, body mass index; WC, waist circumference; WHR, waist-hip-ratio.

**Table S3. The effect of between- and within-family PRS and family history on BMI, WC, and WHR among 2 siblings (n=15,832)**

| Phenotypes | Model | Predictors | Beta | SE | p | $R^2$ (%) |
|---|---|---|---|---|---|---|
| BMI | 1 | Between-family PRS | 0.30 | 0.009 | $< 2.00\times10^{-16}$ | 6.70 |
| | | Within-family PRS | 0.28 | 0.011 | $< 2.00\times10^{-16}$ | 1.91 |
| | 2 | FH | 0.28 | 0.010 | $< 2.00\times10^{-16}$ | 5.07 |
| | | Between-family PRS | 0.24 | 0.010 | $< 2.00\times10^{-16}$ | |
| | | Within-family PRS | 0.27 | 0.014 | $< 2.00\times10^{-16}$ | |
| | 3 | FH | 0.21 | 0.010 | $< 2.00\times10^{-16}$ | 11.72 |
| | | Between-family PRS*FH | 0.03 | 0.011 | $6.13\times10^{-3}$ | |
| | | Within-family PRS*FH | 0.06 | 0.016 | $7.86\times10^{-5}$ | |
| WC | 1 | Between-family PRS | 0.23 | 0.008 | $< 2.00\times10^{-16}$ | 4.10 |
| | | Within-family PRS | 0.23 | 0.010 | $< 2.00\times10^{-16}$ | 1.25 |
| | 2 | FH | 0.16 | 0.007 | $< 2.00\times10^{-16}$ | 3.12 |
| | | Between-family PRS | 0.18 | 0.011 | $< 2.00\times10^{-16}$ | |
| | | Within-family PRS | 0.21 | 0.017 | $< 2.00\times10^{-16}$ | |
| | 3 | FH | 0.13 | 0.007 | $< 2.00\times10^{-16}$ | 7.52 |
| | | Between-family PRS*FH | 0.02 | 0.007 | 0.013 | |
| | | Within-family PRS*FH | 0.02 | 0.011 | 0.054 | |
| WHR | 1 | Between-family PRS | 0.19 | 0.007 | $< 2.00\times10^{-16}$ | 2.61 |
| | | Within-family PRS | 0.17 | 0.011 | $< 2.00\times10^{-16}$ | 0.72 |
| | 2 | FH | 0.11 | 0.007 | $< 2.00\times10^{-16}$ | 1.32 |
| | | Between-family PRS | 0.18 | 0.011 | $< 2.00\times10^{-16}$ | |
| | | Within-family PRS | 0.16 | 0.017 | $< 2.00\times10^{-16}$ | |
| | 3 | FH | 0.09 | 0.006 | $< 2.00\times10^{-16}$ | 4.26 |
| | | Between-family PRS*FH | -0.01 | 0.007 | 0.103 | |
| | | Within-family PRS*FH | 0.01 | 0.011 | 0.350 | |

FH, family history; PRS, polygenic risk score; BMI, body mass index; WC, waist circumference; WHR, waist-hip-ratio. We adjusted age, sex, chips (GSA vs CytoSNP) and 10 principal components in model 0. Model 1: model 0+between-family PRS+within-family PRS. Model 2: model 0+FH. Model 3: model 0+between-family PRS+within-family PRS+FH+between-family PRS*FH+within-family PRS*FH. $R^2$ represented the variance explained by PRS, or QFH, or their combination in addition to model 0.

**Table S4. The effect of between- and within-family PRS and family history on BMI, WC, and WHR among all siblings (n=17,680)**

| Phenotypes | Model | Predictors | Beta | SE | p | $R^2$ (%) |
|---|---|---|---|---|---|---|
| | 1 | Between-family PRS | 0.30 | 0.009 | $< 2.00\times10^{-16}$ | 6.47 |
| | | Within-family PRS | 0.29 | 0.011 | $< 2.00\times10^{-16}$ | 2.22 |
| | 2 | FH | 0.27 | 0.010 | $< 2.00\times10^{-16}$ | 4.88 |
| BMI | | Between-family PRS | 0.24 | 0.010 | $< 2.00\times10^{-16}$ | |
| | | Within-family PRS | 0.27 | 0.014 | $< 2.00\times10^{-16}$ | |
| | 3 | FH | 0.20 | 0.010 | $< 2.00\times10^{-16}$ | 11.73 |
| | | Between-family PRS*FH | 0.03 | 0.011 | 0.018 | |
| | | Within-family PRS*FH | 0.07 | 0.015 | $6.46\times10^{-6}$ | |
| | 1 | Between-family PRS | 0.23 | 0.008 | $< 2.00\times10^{-16}$ | 3.95 |
| | | Within-family PRS | 0.24 | 0.010 | $< 2.00\times10^{-16}$ | 1.49 |
| | 2 | FH | 0.15 | 0.007 | $< 2.00\times10^{-16}$ | 2.86 |
| WC | | Between-family PRS | 0.19 | 0.011 | $< 2.00\times10^{-16}$ | |
| | | Within-family PRS | 0.20 | 0.016 | $< 2.00\times10^{-16}$ | |
| | 3 | FH | 0.12 | 0.007 | $< 2.00\times10^{-16}$ | 7.48 |
| | | Between-family PRS*FH | 0.01 | 0.007 | 0.071 | |
| | | Within-family PRS*FH | 0.04 | 0.010 | $3.86\times10^{-4}$ | |
| | 1 | Between-family PRS | 0.19 | 0.007 | $< 2.00\times10^{-16}$ | 2.56 |
| | | Within-family PRS | 0.17 | 0.010 | $< 2.00\times10^{-16}$ | 0.84 |
| | 2 | FH | 0.10 | 0.006 | $< 2.00\times10^{-16}$ | 1.24 |
| WHR | | Between-family PRS | 0.19 | 0.011 | $< 2.00\times10^{-16}$ | |
| | | Within-family PRS | 0.16 | 0.016 | $< 2.00\times10^{-16}$ | |
| | 3 | FH | 0.09 | 0.006 | $< 2.00\times10^{-16}$ | 4.33 |
| | | Between-family PRS*FH | -0.02 | 0.007 | 0.024 | |
| | | Within-family PRS*FH | 0.02 | 0.010 | 0.045 | |

FH, family history; PRS, polygenic risk score; BMI, body mass index; WC, waist circumference; WHR, waist-hip-ratio. We adjusted age, sex, chips (GSA vs CytoSNP) and 10 principal components in model 0. Model 1: model 0+between-family PRS+within-family PRS. Model 2: model 0+FH. Model 3: model 0+between-family PRS+within-family PRS+FH+between-family PRS*FH+within-family PRS*FH. $R^2$ represented the variance explained by PRS, or QFH, or their combination in addition to model 0.

The effect sizes and variance explained by between-family PRS and within-family PRS formula:

Part1:

$$y = b_1(x - \bar{x}) + b_2\,\bar{x}$$

$$\bar{x} = \frac{1}{2}(x_1 + x_2)$$

$$v(x - \bar{x}) = v\left[x - \frac{1}{2}(x_1 + x_2)\right]$$

$$= v\left[x_1 - \frac{1}{2}(x_1) - \frac{1}{2}(x_2)\right]$$

$$= v\left(\frac{1}{2}x_1 - \frac{1}{2}x_2\right)$$

$$= \frac{1}{4}[v(x_1) + v(x_2) - 2cov]$$

$$= \frac{1}{4}\left[2v(x) - 2 \times \frac{1}{2}v(x)\right]$$

$$= \frac{1}{4}v(x)$$

$$v(\bar{x}) = \frac{3}{4}v(x)$$

where y is the outcome, $x_1$ and $x_2$ are the PRS of first and second siblings, $\bar{x}$ is the mean PRS of two siblings, representing between family effect, $(x - \bar{x})$ represents within-family PRS effect. The within-siblings effect $b_1$ represents the expected change given a one-unit change in the difference between the individual PRS and the average PRS of the siblings. The between-siblings effect $b_2$ represents the expected change in the outcome y given a one-unit change in the average of the family PRS. $v(x)$ represents the variance of outcome explained by PRS. $v(x - \bar{x})$ represents the variance of outcome explained by the within-family PRS effects. $v(\bar{x})$ represents the variance explained by the between-family PRS effects.

Part 2:

$$cov[(x_1 - \bar{x}), \bar{x}] = cov(x_1, \bar{x}) - v(\bar{x}) = cov\left[x_1, \frac{1}{2}(x_1 + x_2)\right] - v(\bar{x}) = \frac{1}{2}v(x) + \frac{1}{4}v(x) - \frac{3}{4}v(x) = 0$$

$$b = \begin{bmatrix} \frac{1}{4}v(x) & 0 \\ & \frac{3}{4}v(x) \end{bmatrix} \begin{bmatrix} \frac{1}{4} \\ \frac{3}{4} \end{bmatrix} cov(y, x)$$

$$cov(y, x_1 - \bar{x}) = \frac{1}{2}cov(y, x_1 - x_2) = \frac{1}{2}\left[cov(y, x) - \frac{1}{2}cov(y, x)\right] = \frac{1}{4}cov(y, x)$$

$$cov(y, \bar{x}) = \frac{1}{2}cov(y, x_1 + x_2) = \frac{3}{4}cov(y, x)$$

where $cov[(x_1 - \bar{x}), \bar{x}]$ represents the correlation between within-family PRS $(x_1 - \bar{x})$ and between-family PRS $(\bar{x})$, and this correlation is zero. $cov(y, x)$ denotes the correlation between the outcome and PRS in general, while $cov(y, x_1 - \bar{x})$ represents the correlation specifically between outcome and within-family PRS, and $cov(y, \bar{x})$ represents the correlation between the outcome and between-family PRS.

Part 3:

For effect size of between- and within-family PRS,

$$\frac{cov(x,y)}{v(x)} \begin{bmatrix} \frac{1}{4} & \\ & \frac{3}{4} \end{bmatrix}^{-1} \begin{bmatrix} \frac{1}{4} \\ \frac{3}{4} \end{bmatrix} = \beta \begin{bmatrix} 4 & 0 \\ 0 & \frac{4}{3} \end{bmatrix} \begin{bmatrix} \frac{1}{4} \\ \frac{3}{4} \end{bmatrix} = \beta \begin{bmatrix} 1 \\ 1 \end{bmatrix}$$

Therefore, $\beta_B = \beta_W$

For the variance explained by within-family PRS,

$$R^2 = \frac{cov(y, x_1 - \bar{x})^2}{v(x - \bar{x})v(y)} = \frac{\frac{1}{16}cov(y,x)^2}{\frac{1}{4}v(x)v(y)} = \frac{1}{4}R^2$$

For the variance explained by between-family PRS,

$$R^2 = \frac{cov(y, \bar{x})^2}{v(\bar{x})v(y)} = \frac{(\frac{3}{4})^2 cov(y,x)^2}{\frac{3}{4}v(x)v(y)} = \frac{3}{4}R^2$$

In theory, between-family PRS explained three times the variance of outcome compared to within-family PRS.

**UMCG Genetics Lifelines Initiative (UGLI) group author**

LifeLines Cohort Study

Raul Aguirre-Gamboa (1), Patrick Deelen (1), Lude Franke (1), Jan A Kuivenhoven (2), Esteban A Lopera Maya (1), Ilja M Nolte (3), Serena Sanna (1), Harold Snieder (3), Morris A Swertz (1), Peter M. Visscher (3,4), Judith M Vonk (3), Cisca Wijmenga (1)

(1) Department of Genetics, University of Groningen, University Medical Center Groningen, The Netherlands
(2) Department of Pediatrics, University of Groningen, University Medical Center Groningen, The Netherlands
(3) Department of Epidemiology, University of Groningen, University Medical Center Groningen, The Netherlands
(4) Institute for Molecular Bioscience, The University of Queensland, Brisbane, Queensland, Australia.

**Acknowledgements**

Lifelines Cohort Study

The Lifelines Biobank initiative has been made possible by funding from the Dutch Ministry of Health, Welfare and Sport, the Dutch Ministry of Economic Affairs, the University Medical Center Groningen (UMCG the Netherlands), University of Groningen and the Northern Provinces of the Netherlands. The generation and management of GWAS genotype data for the Lifelines Cohort Study is supported by the UMCG Genetics Lifelines Initiative (UGLI). UGLI is partly supported by a Spinoza Grant from NWO, awarded to Cisca Wijmenga. The authors wish to acknowledge the services of the Lifelines Cohort Study, the contributing research centers delivering data to Lifelines, and all the study participants.
